# Optimizing Mobile Health Clinic Placement via Geospatial Modeling

**DOI:** 10.64898/2025.12.29.25342286

**Authors:** Shakhawat H. Tanim, David White, Brian Witrick, Lior Rennert

## Abstract

**Objectives:** Mobile health clinics (MHCs) provide flexible, community-based care to underserved populations facing geographic and socioeconomic barriers. Maximizing coverage enables MHCs to reach more individuals, improve preventive and continuous care, and reduce health disparities. However, few strategies exist to guide placement and routing decisions. We present a framework to increase MHC utilization by optimizing service coverage.

**Study Design:** This is a retrospective study.

**Methods:** We analyzed MHC deployments for Hepatitis C Virus (HCV) screening and treatment from a local health system in South Carolina. We used a location-allocation model to identify potential MHC placement sites that maximized the number of uninsured residents within a 5-minute drive or 10-minute walk. Demand was represented by block centroids weighted by the size of the uninsured population. We compared service area population, defined as the size of the target population within driving or walking distance, for model-proposed sites with coverage from previous MHC deployments. We fit negative binomial mixed effects models to evaluate the association between service area population and MHC utilization.

**Results:** Optimized placements can nearly double population coverage, expanding access to uninsured residents within practical travel distances by 90% for driving and 135% for walking—without requiring additional vehicles or resources. This approach also substantially reduces redundant service areas while shortening average travel times. Results show that small geographic shifts can yield significant improvements. In rural regions, greater geographic coverage is significantly associated with higher MHC utilization for HCV screening (drive p=.0037; walk p=.0095). We applied this framework with local health partners to guide real-world MHC deployment in South Carolina.

**Conclusions:** This framework connects spatial analytics to service delivery, offering a replicable, operationally ready tool adaptable to various travel modes, site types, and disease contexts. It supports strategic placement in high-need locations by reducing travel time and service redundancy and ultimately improving health outcomes in medically underserved populations.

## Background

Mobile health clinics (MHCs) play an important role in the healthcare system, delivering essential care to most underserved populations across countries such as the U.S., Canada, the UK, France, Norway, Brazil, Egypt, Iraq, India, South Africa, and more.^1–4^ These vehicle-based clinics, staffed by healthcare professionals and equipped with medical supplies, directly address the fundamental barriers that prevent vulnerable populations from accessing care: transportation limitations, geographic isolation, and healthcare system complexity.^1,2,5^ By bringing preventive care and chronic disease management directly to medically underserved communities,^1,6–8^ MHCs not only improve health outcomes and reduce costs,^6,9^ but also alleviate the burden on emergency departments.^10^ The COVID-19 pandemic further demonstrated their importance, as MHCs expanded their functions in managing public health crises.^11,12^

Yet despite their demonstrated impact, MHCs face a fundamental challenge that undermines their potential to reach medically underserved: the absence of strategic, systematic methods for determining where these resources should be deployed. Currently, many MHC programs rely on geographic and logistic convenience^13^ rather than systematic approaches to site selection. Since site placement influences accessibility of MHCs, poor placement can lead to missed opportunities to improve utilization. For example, data from our recent study on MHC deployment for HCV screening and treatment, demonstrated substantial variation in utilization, ranging from 1 to 43 patients screened across 98 sites’ deployment, with an average of 4.81 [SD = 5.89] screened per visit.^7^ This wide variability strongly suggests that site location is a key driver of patient engagement and that a systematic approach for optimum placement could consistently yield higher utilization. Compounding the issue, patient surveys from these deployments reveal that more than half of the patients had to use non-car modes like walking or transit to reach the MHCs, showing that even MHCs can remain out of reach for some people with transportation barriers, particularly for uninsured individuals who often rely on public transit or walking.^14^

To address these limitations, the lack of a data-based optimal placement strategy, the high variability of utilization, and geographic and transportation accessibility, geospatial methods can be used for facility placement decisions. Geospatial location-allocation models, which strategically determine optimal facility locations,^15^ have been used for fixed-site facilities to maximize service coverage,^16^ promoting accessibility,^17^ and improving operational efficiency.^18^ However, mobile and fixed healthcare delivery serve fundamentally different purposes and operate under distinct constraints. Fixed-site optimization typically prioritizes maximizing patient volume in densely populated urban centers, an objective that conflicts with the core mission of MHCs, which is to reach medically underserved communities that may be geographically dispersed (e.g., rural regions) or require targeted interventions. Additionally, traditional fixed-site models rely on static demographic data that fail to capture the dynamic community needs and real-world service utilization patterns essential for responsive mobile health planning.

In sum, despite their clear benefits, MHCs are typically deployed without a systematic, data-driven placement strategy. To address these gaps, we developed a geospatial framework specifically designed for optimizing MHC placement decisions through increasing service area population. Our approach bridges the gap between traditional facility planning methods and the unique requirements of mobile health delivery, offering healthcare decision-makers a systematic, evidence-based tool for strategic resource allocation. By incorporating empirical deployment data and real-world utilization patterns, this framework enables more effective targeting of mobile health resources, ultimately expanding access to care for the communities that need it most.

## Methods

### Study Setting and Design

We used operational data from Clemson Rural Health (CRH)’s MHC initiative for Hepatitis C Virus (HCV) screening and treatment to develop and evaluate a framework for optimizing MHC placement to reach uninsured populations.^7^ CRH has deployed MHCs across South Carolina for hepatitis C virus (HCV) screening and treatment in underserved communities. Analyses were restricted to ZIP codes in the Upstate region with prior MHC activity, enabling direct comparison between observed sites and sites optimized by our framework. This study was approved by the Institutional Review Board of Clemson University (Protocol #IRB2022-0150) and by the Prisma Health institutional review board (IRB Pro 00106348).

### Data Sources

#### Operational Data

We analyzed de-identified records from CRH’s HCV-focused MHC program (May 24, 2021–November 13, 2024): 261 deployment events across 48 unique locations, documenting services provided to 1,166 individuals. Each record included site name, address, site classification, service date, and number of patients served. To ensure our analysis reflected community accessibility, we excluded deployment sites serving captive populations (detention centers and residential behavioral health facilities).

#### Population and Geographic Data

Population-level insurance coverage data were obtained from the American Community Survey, accessed at the census block group level.^19^ We disaggregated these data to individual census blocks using Geographic Correspondence Engine,^20^ enabling fine-scale spatial analysis of uninsured populations. Urban-rural classifications followed the 2020 U.S. Census Bureau definitions.^21^

#### Transportation Network and Candidate Facility Data

Road and pedestrian networks were sourced from Esri StreetMap Premium.^22^ Candidate facility locations, representing the potential MHC deployment sites, were identified through ArcGIS Business Analyst,^23^ with selections restricted to site types compatible with observed MHC site types.

### Procedures

#### Location-Allocation Optimization Modeling

We employed location-allocation modeling to identify optimal MHC sites that would maximize coverage of uninsured populations within realistic travel times. This approach simultaneously determines the best facility locations and assigns populations to these facilities based on accessibility. For each of the 20 ZIP codes, we maintained the same number of sites as observed sites but optimized their locations. We modeled two transportation scenarios: 5-minute driving and 10-minute walking thresholds, reflecting realistic access patterns.^24,25^ Census block centroids served as demand points, weighted by the number of uninsured residents. The objective function maximized covered population, assuming equal capacity across sites and network travel times along actual roads.

#### Accessibility Assessment and Comparative Analysis

We generated network-based service areas around both observed and proposed MHC sites using 5-minute drive and 10-minute walk thresholds. By overlaying these with census block centroids, we calculated the number of uninsured residents within reach, which defined service area coverage population, and compared ZIP based coverage between observed deployments and the proposed configuration.

### Statistical analysis

To test whether greater geographic coverage translates to higher utilization, we modeled patients served per deployment using a negative binomial mixed-effects model (GLMM). The principal predictor was the number of uninsured residents within the site’s service area, with urban–rural status and their interaction included to assess heterogeneity by geographic context.

### Software and reproducibility

Geospatial analyses were performed in ArcGIS Pro 3.4 (Network Analyst; StreetMap Premium; ArcGIS Business Analyst). Statistical analyses were conducted in R 4.3.1.

## Results

### Location Allocation

This section first presents detailed findings for selected ZIP codes to illustrate how spatial optimization improves placement for both single-site and multi-site scenarios. It then provides a combined regional summary for the Upstate area based on service area metrics. In this context, observed sites are previous MHC deployments, proposed sites are model-optimized locations, and candidate sites are all potential locations considered.

#### Single Site Scenario: Minimizing Driving Time

In a predominantly rural ZIP code with one prior MHC deployment, the model proposes a single MHC location from 58 candidate sites (Figure 1). The proposed site lies centrally within a dense cluster of uninsured residents whereas the observed site is more peripheral. The proposed site captures 257 census blocks within a 5-minute drive and offers an average travel time of 3.7 minutes per person, indicating easy spatial accessibility. The service area analysis reveals that the proposed site encompasses 783 uninsured individuals within a 5-minute drive, representing a 32-fold increase compared to the 23 individuals within the service area of the observed site for the CRH HCV screening program.

**Figure 1:**
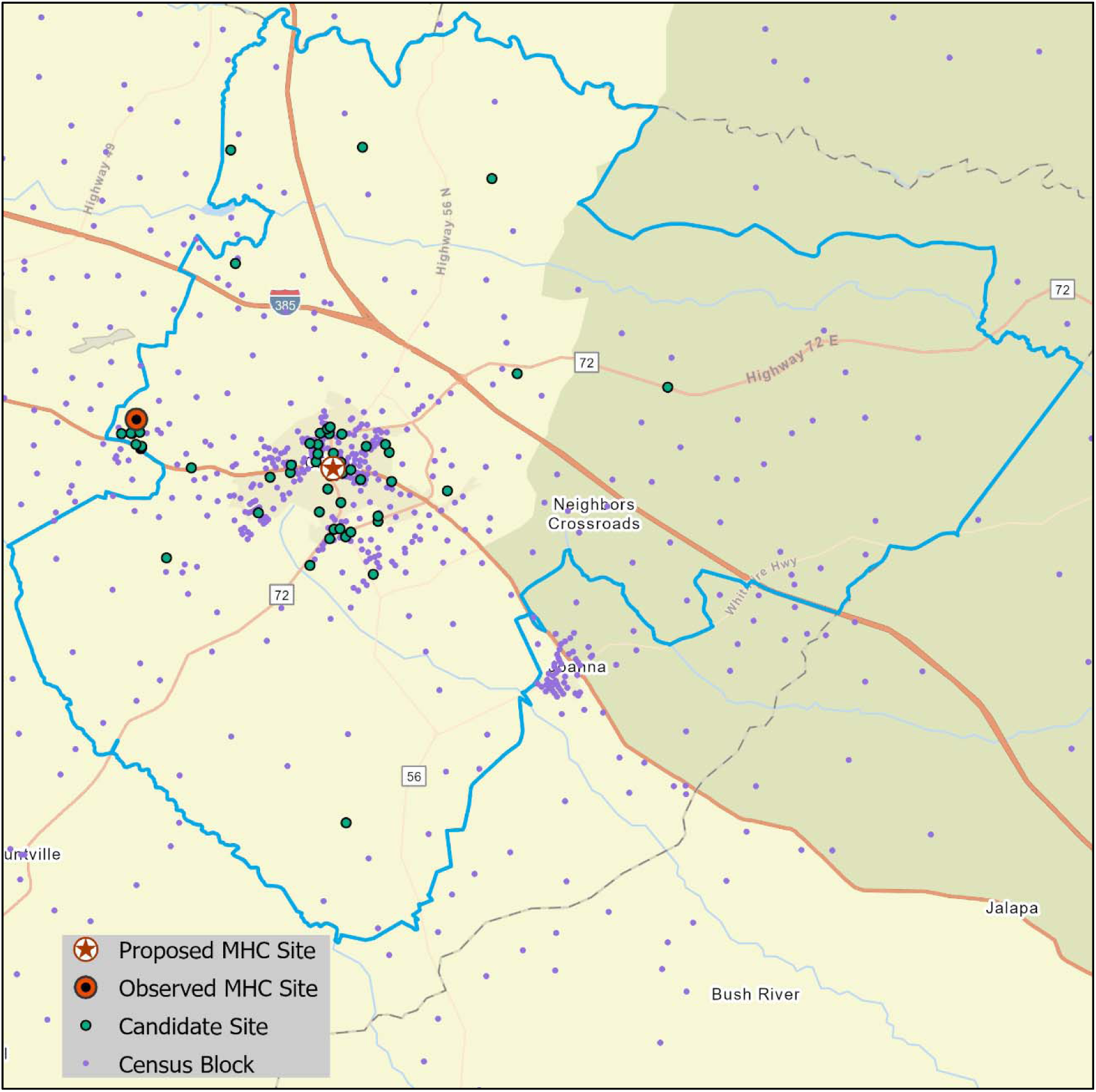
Location allocation for a single-site scenario for driving time, showing the spatial distribution of the proposed site, observed site, candidate sites, and census block centroids in a rural ZIP code.

#### Multi-site Scenario: Minimizing Driving Time

For a dense urban ZIP code within a metropolitan setting with three observed MHC placements, the model optimized the same number of proposed sites from 140 candidate sites. Figure 2 shows the spatial distribution of these sites. The proposed configuration covers 1,410 census blocks within the 5-minute drive threshold, achieving an average travel time of 4.16 minutes per person. The proposed sites offer clear advantages over the observed sites in terms of spatial coverage. While the observed sites are concentrated primarily in the western portion of the area, the proposed sites extend service reach across the entire area. For instance, the proposed site in the northwest corner reaches uninsured residents who were previously unserved. Service area analysis reveals that observed MHC sites could serve unique 2,832 uninsured individuals, while the proposed sites cover 7,882, an increase of 5,050 individuals, or a 179% improvement.

**Figure 2:**
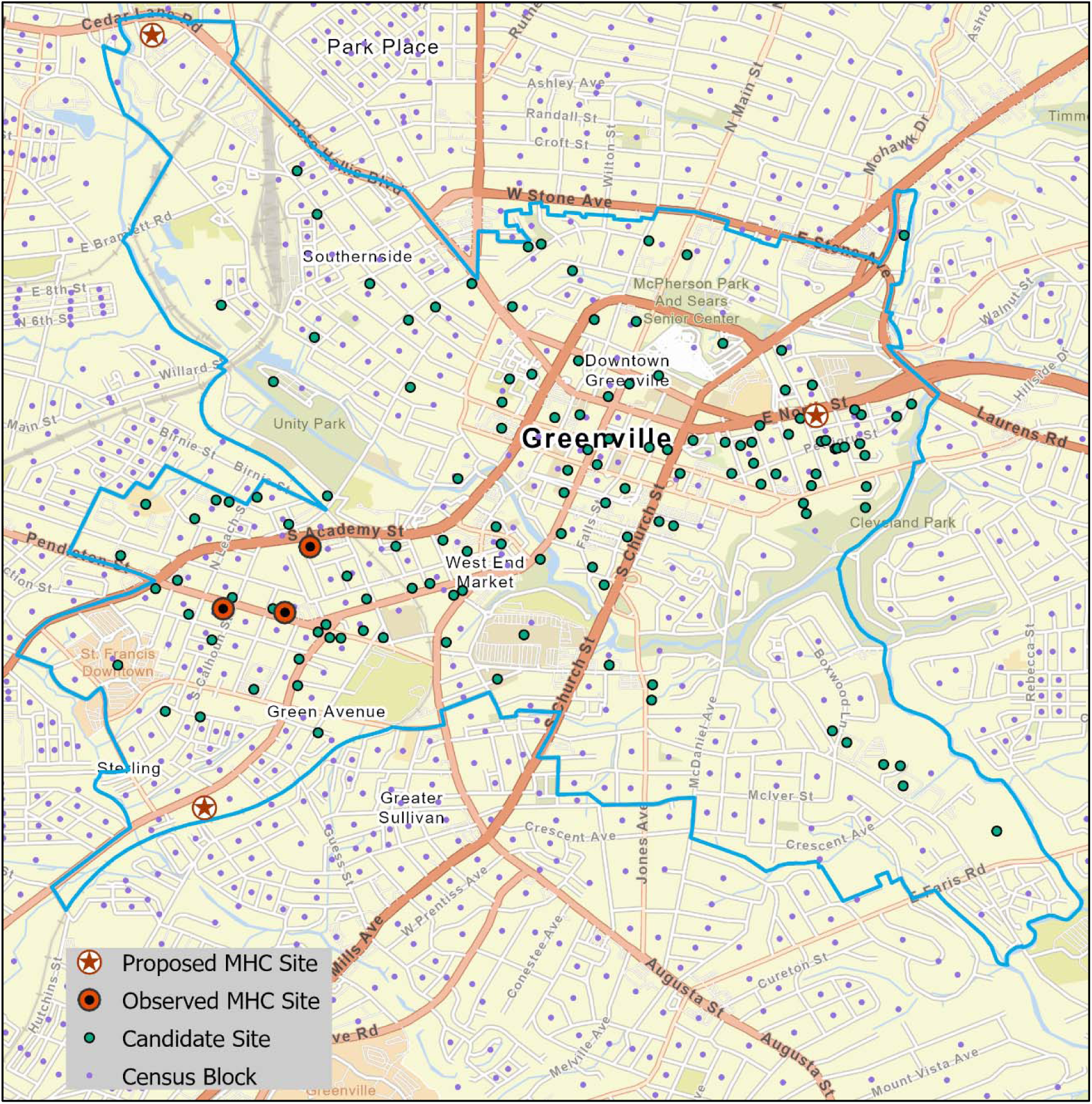
Location allocation for a multi-site scenario for driving time, showing the spatial distribution of three proposed sites, three observed sites, candidate sites, and census block centroids in an urban ZIP code.

Findings for this area show how optimized placement reduces spatial overlap and expands access. Overall, four ZIP codes had multiple sites with overlapping service areas i.e., uninsured residents in a site’s service area who also lie within areas covered by other sites in the same ZIP. In the driving scenario, the proposed sites reduce the uninsured population overlap by 87%, 95%, 96%, and 100%, effectively eliminating most duplication.

Additional ZIP code-level analyses for walking scenarios are presented in the appendix (p 1-2).

#### Regional Analysis: Comparing Observed Service Areas to Model-Selected Service Areas

Figure 3 (panels a–b) shows the distribution of observed and proposed MHC sites with census blocks across the Upstate region under driving access and walking access scenarios. These composite maps aggregate all ZIP codes with previous MHC deployments. While the regional scale limits visibility of individual site movements, detailed ZIP-level maps (Figures 2-3, appendix p 1-2) demonstrate specific placement improvements and coverage impacts.

**Figure 3.**
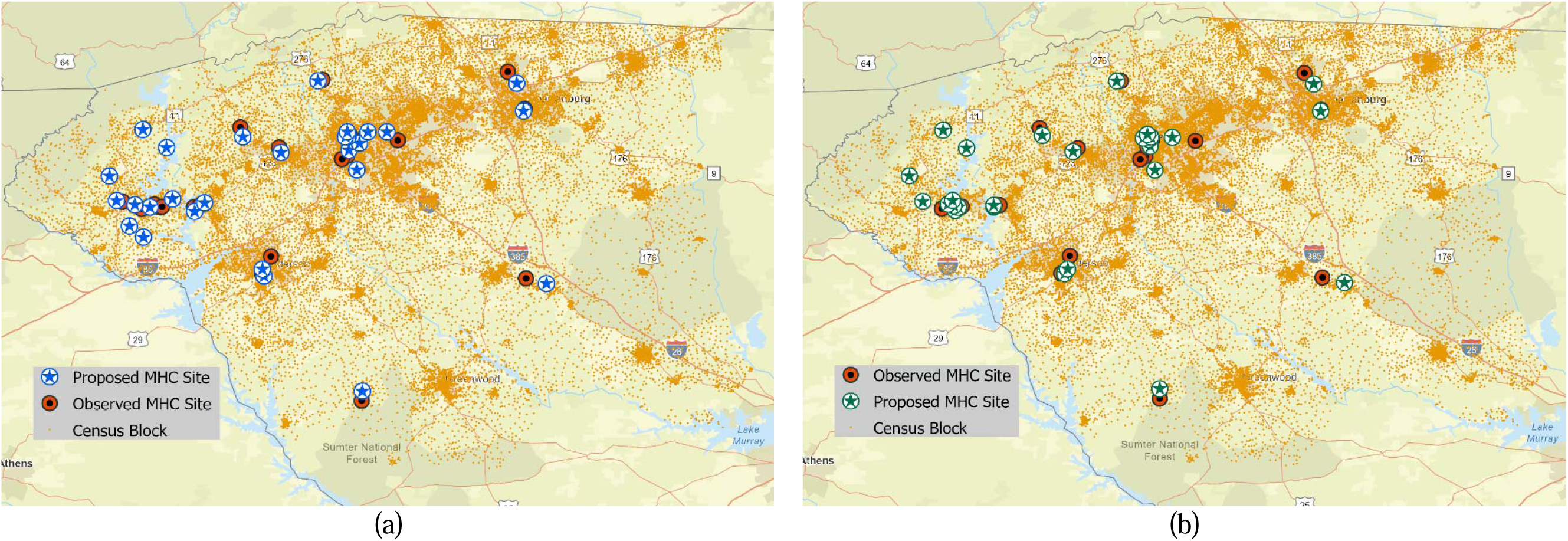
Spatial distribution of observed and proposed MHC sites with census blocks (demand points) across the Upstate South Carolina region under (a) driving access (5-minute) and (b) walking access (10-minute) scenarios.

To compare how the reach of proposed sites improved compared to observed sites, we analyzed the service area population of observed versus proposed MHC site configurations across ZIP codes for both driving and walking access. Table 1 summarizes the number of uninsured individuals served under each configuration.

**Table 1:** Comparative analysis of MHC service coverage under driving and walking access scenarios across ZIP codes for unique observed and proposed site placements, with corresponding uninsured population reach and absolute and percentage improvements.

| ZIP Code | Site Count | Uninsured population in driving service area |  | Difference | Uninsured population in walk service area |  | Difference | Rural/Urban |
| --- | --- | --- | --- | --- | --- | --- | --- | --- |
|  |  | Observed MHC sites | Proposed MHC sites |  | Observed MHC sites | Proposed MHC sites |  |  |
| 29306 | 1 | 1812 | 1890 | 78 (4%) | 116 | 242 | 126 (109%) | Rural |
| 29316 | 1 | 417 | 1036 | 620 (149%) | 2 | 114 | 112 (4,620%) | Urban |
| 29325 | 1 | 23 | 783 | 760 (3,294%) | 1* | 101 | 100 (10,032%) | Rural |
| 29601 | 3 | 2832 | 7799 | 4,967 (175%) | 355 | 842 | 487 (137%) | Urban |
| 29605 | 1 | 1751 | 2295 | 544 (31%) | 262 | 292 | 30 (11%) | Urban |
| 29609 | 2 | 4844 | 6763 | 1,918 (40%) | 706 | 891 | 185 (26%) | Urban |
| 29611 | 1 | 1786 | 6283 | 4,497 (252%) | 6 | 974 | 968 (16,187%) | Urban |
| 29615 | 1 | 563 | 2296 | 1,733 (308%) | 10 | 240 | 230 (2,212%) | Urban |
| 29620 | 1 | 119 | 504 | 385 (324%) | 1* | 89 | 88 (8,834%) | Rural |
| 29621 | 1 | 352 | 2217 | 1,865 (530%) | 14 | 309 | 296 (2,188%) | Rural |
| 29624 | 1 | 2297 | 2718 | 421 (18%) | 183 | 326 | 142 (78%) | Rural |
| 29631 | 1 | 427 | 1175 | 748 (175%) | 79 | 194 | 115 (145%) | Urban |
| 29634 | 1 | 403 | 501 | 98 (24%) | 320 | 320 | 0 (0%) | Urban |
| 29640 | 1 | 793 | 1054 | 260 (33%) | 70 | 157 | 87 (125%) | Rural |
| 29661 | 1 | 342 | 378 | 36 (11%) | 1* | 23 | 22 (2,220%) | Rural |
| 29671 | 1 | 327 | 609 | 282 (86%) | 6 | 118 | 112 (1,863%) | Rural |
| 29676 | 2 | 51 | 59 | 8 (16%) | 2 | 8 | 6 (294%) | Rural |
| 29678 | 5 | 1348 | 1923 | 575 (43%) | 248 | 582 | 334 (135%) | Rural |
| 29691 | 1 | 1279 | 1279 | 0 (0%) | 230 | 350 | 120 (52%) | Rural |
| 29693 | 1 | 277 | 396 | 119 (43%) | 28 | 28 | 0 (0%) | Rural |
| **Total** | 28 | 22043 | 41957 | 19,914 (90%) | 2641 | 6202 | 3,561 (135%) | - |

*\* For ZIP codes with zero observed service area population, a minimum value of 1 was used to calculate percentage increases.*

The proposed driving configuration served 41,957 uninsured individuals, a 90% increase over the 22,043 served by observed sites. Coverage of service area population improved in 19 of 20 ZIP codes; ZIP 29691 remained unchanged due to identical site placement. The highest relative gain occurred in rural ZIP 29325, driven by a very low baseline, while urban ZIP 29601 showed the greatest absolute gain. Urban areas, such as 29601, 29611, and 29609, contributed the most to total increases due to higher population densities, while rural ZIPs, including 29325, 29621, and 29620, saw large relative gains from small baselines.

In the walking scenario, service area population increased from 2,641 to 6,202 individuals, a 135% increase. ZIP codes 29634 and 29693 showed no change due to unchanged site locations. The largest gain in both absolute and relative terms occurred in urban ZIP 29611 where the original baseline was very low. ZIPs with no observed service area population were assigned a minimum of 1 in percentage calculations. Excluding those, urban ZIPs such as 29611, 29316, and 29615 still demonstrated substantial relative improvements, while absolute gains remained distributed across both urban and rural ZIPs. Overall, urban areas captured most of the absolute growth because of larger baseline populations whereas rural areas, however, experienced the steepest relative improvements particularly for walking access (appendix p 3).

### Association Between Observed Service Area and MHC Utilization

Analysis of 98 MHC deployments for HCV screening across South Carolina revealed a positive association between service area population and utilization (appendix p 4). However, the association between service area population and utilization was only statistically significant in rural areas (5-min drive: p=.0037; 10-min walk: p=.0095). While urban sites were associated with an overall greater increase in utilization compared to rural areas (drive: p=.0198; walk: p=.0139), we did not observe a significant effect of service area population within urban sites (drive: p = 0.915; walk: p = 0.108).

### *Case* S*tudy*: Real-World Implementation

The framework’s transition from theoretical model to operational tool is exemplified through three deployment cases with CRH and Prisma Health, each revealing distinct insights about spatially optimized healthcare delivery. These applications demonstrate not merely the framework’s versatility, but its capacity to transform abstract optimization into actionable deployment strategies across diverse epidemiological contexts.

Case Study 1 was implemented for County X in South Carolina, which we deidentified to avoid stigmatization. This county faces limited healthcare infrastructure with only one hospital, making it a candidate county for MHC services. Currently, CRH does not operate in County X. In collaboration with CRH, we implemented our framework to identify candidate sites within county X for potential expansion of MHC services. We focused on ZIP code Y within the county, as this was identified as high-priority area due to disproportionately high rates of HCV-related hospitalizations and mortality.

Using our framework, we evaluated 150 candidate sites in ZIP code Y, representing a diverse mix of community-accessible locations, including churches, businesses, and public service centers. From this pool, 20 optimal sites were identified for potential MHC deployment (Figure 4). The model prioritized uninsured adults aged 35 to 65, an at-risk group for HCV.^7,26^ The entire process, from county and ZIP code selection to site optimization, integrated demographic, epidemiological, and transportation data to ensure maximum coverage for target underserved populations.

**Figure 4:**
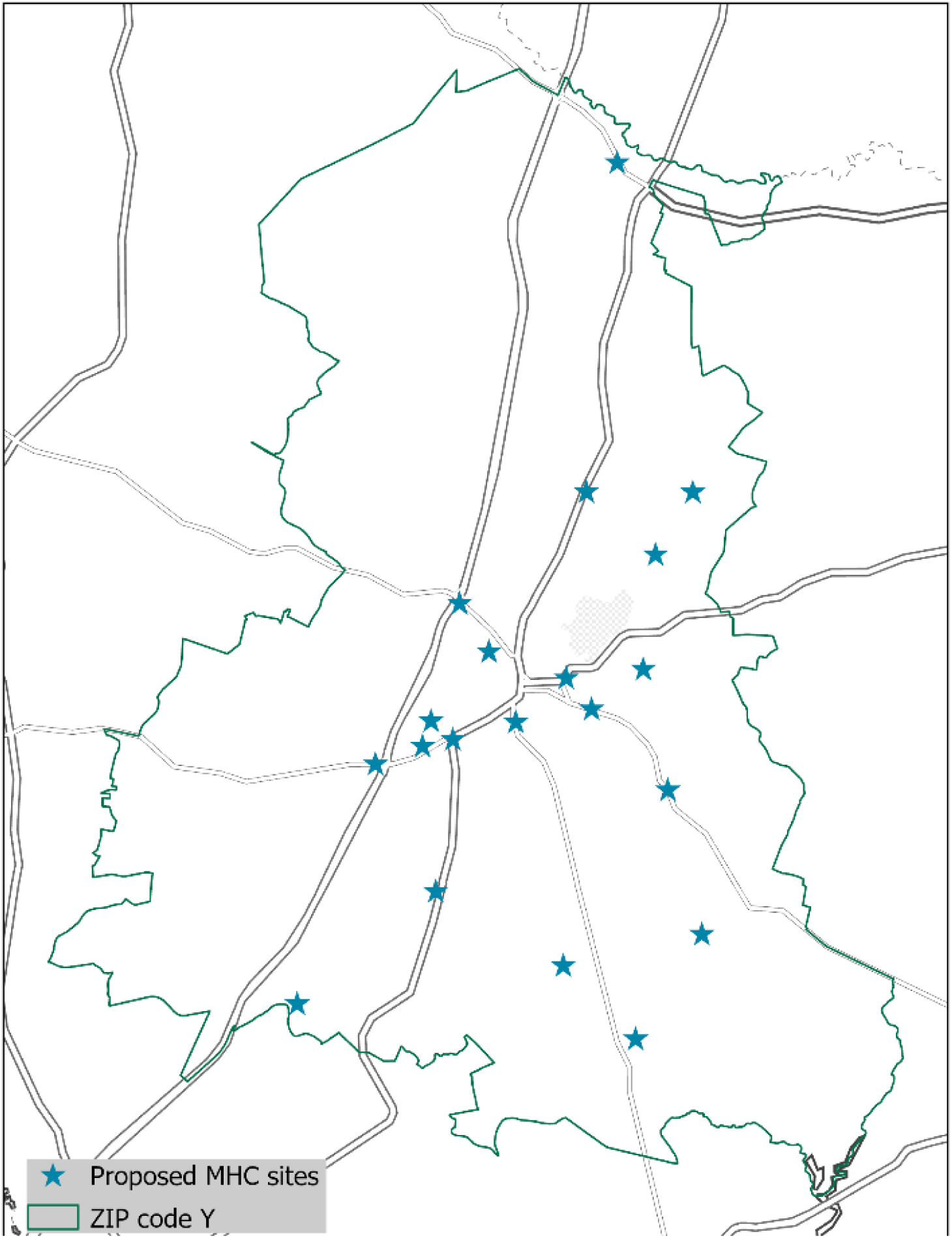
Distribution of 20 proposed MHC sites in X County, South Carolina, identified to support future deployment planning by Clemson Rural Health.

The selected sites demonstrated strong spatial coverage for the target population residing within a 5-minute drive or 10-minute walk service area, to maximize population reach. We ranked sites by the number of uninsured adults aged 35–65 covered (appendix p 5). Collectively, the 20 optimal sites for 5-minute drive service areas, provided potential coverage for 45,652 individuals, including 4,335 uninsured adults aged 35–65. The top nine sites each cover more than 250 uninsured adults within a 5-minute drive.

In another application (Case Study 2), we applied this model to prioritize MHC deployments for respiratory virus screening at sites where CRH and Prisma Health currently operate (funded by the CDC Center for Forecasting and Outbreak Analytics). We first identified 11 priority ZIP codes based on the highest mortality rates from COVID-19, influenza, and RSV. Within these high-risk areas, we evaluated 32 MHC sites operated by our partners, CRH and Prisma Health. We ranked each site based on its coverage of uninsured adults aged 35–65 within both 10-minute drive and 10-minute walk service areas (appendix p 6). Collectively, these sites covered 63,218 uninsured adults aged 35-65 within a 10-minute drive, with the top site (deidentified S-1) serving 4,846 individuals. Although S-1 showed the highest reach for both driving and walking, this pattern was not consistent across all sites. Some sites demonstrated high coverage only for driving access, while others excelled primarily in walking access. Additionally, a ZIP code-focused approach, prioritizing ZIP codes with high mortality rates, can help identify optimal sites within those ZIP codes. For example, among the three sites located in Zip Code Z-5 (deidentified), which has one of the highest death rates, site S-3 provides the greatest coverage.

In the third application (Case Study 3), funded by the National Institute on Drug Abuse (NIDA) of the National Institutes of Health (NIH) for the delivery of medications against opioid use disorder (MOUD) via MHC, we selected three sites from six candidate sites (appendix p 7). The need to deploy MHCs within proximity to 340B pharmacies, so patients could easily fill prescriptions, limited the number of candidate sites for this case. We evaluated the candidate sites, calculating combined service area populations within both a 10-minute walk and a 5-minute drive for uninsured adults aged 35–64. Sites S-1, S-2, and S-3 (deidentified) were identified as the optimal three-site configuration, collectively serving 1,550 uninsured adults aged 35–64. This configuration maximizes reach to the target population and minimizes redundancy from overlapping service areas.

## Discussion

This study demonstrates how geospatial location-allocation analysis, combined with real-world operational data, transforms MHC placement from convenience-based practices into a systematic, data-driven, and evidence-based process. A central finding is that reallocating existing sites unlocks substantial latent capacity. Using the same 28 clinics, optimized placement increased the uninsured population within reach by 90% for 5-minute driving and 135% for 10-minute walking. These gains arose from shifting clinics toward demand clusters, reducing overlap among service areas, and minimizing travel times. In one example ZIP Code, moving a single clinic from a peripheral to a central location increased the number of uninsured individuals serviced within a 5-minute drive from 23 to 783 and within a 10-minute walk from 0 to 101, illustrating how small geographic shifts can yield significant improvements.^27^ Large increases in dense urban ZIP codes, together with substantial gains in rural areas, indicate that the approach advances coverage of the medically underserved population in both settings.

Findings suggest that expanding the service area’s population and improving accessibility can be achieved simultaneously, depending on the context. In multi-site urban ZIP codes, dispersing clinics reduced clustering, extended coverage to previously underserved areas, and increased reach to the target population. In rural ZIP codes, where populations are dispersed and baseline access is limited, spatial adjustments substantially increased the reach of the target population.

Driving and walking-based optimization reveal different realities. Driving scenarios expand overall reach, whereas walking scenarios expose pedestrian reachable areas connected with walkable paths or sidewalks.^28^ The consistently larger relative gains under the walking configuration indicate that future placement has opportunities to increase accessibility for pedestrians. Potentially, this neighborhood-scale placement of health care facilities is important for populations reliant on proximity due to limited mobility or transportation options,^29^ including households without vehicles.^30^ Consequently, multimodal planning is essential because optimizing for a single mode may appear efficient in aggregate, yet leaves access gaps for those who do not have access to a car.^31^

Beyond increasing coverage and accessibility, the framework supports more patient-centered planning and evaluation. By producing block-level travel times, it provides operational benchmarks such as the average travel time or high percentile travel times that can be used to evaluate the gains. This framework also enables scenario testing, for example, reallocating sites across days or co-locating with community partners, before committing full resources.

The framework’s transition from analytic proof-of-concept to an operational decision-support tool within large health systems is illustrated through collaborative deployments with Clemson Rural Health and Prisma Health, where it now directly informs site selection for MHCs addressing screening and treatment for HCV, respiratory viruses, and opioid use disorder. Through these deployments, our MHC partners are already using the framework to prioritize high-need ZIP codes, identify feasible deployment sites, and coordinate logistics, demonstrating its practical impact, scalability, and immediate policy relevance.

Linking the target population with observed utilization confirms the approach’s practical relevance. A positive association between the uninsured population within service areas and MHC utilization for HCV screening suggests that expanding geographic access is associated with greater uptake in rural areas. Yet in urban settings the effect is attenuated, indicating that non-spatial determinants such as community partner networks,^7,32^ outreach,^33^ trust,^34^ and cultural relevance^35^ can mediate how well access converts into use. However, this is based on a single site setting for a specific disease and therefore should be interpreted with caution.

All these findings have direct implications for health policy and system management. Before expanding MHC fleets, health systems should ensure that resources are optimally deployed. The potential to nearly double population coverage without additional investment offers an immediate opportunity to extend reach and reduce spatial barriers to healthcare. By integrating fine-scale geospatial analytics with real-world implementation, this framework directly supports efforts to advance health equity and system efficiency. Embedding this framework in routine planning cycles, supported by investment in GIS infrastructure, high-resolution data, routinely updated spatial layers, and local capacity, can improve future placement decisions. These improvements to mobile health clinic planning strengthen pandemic preparedness^36^ and enhance emergency response to more frequent and severe climate-related events.^37^ Additionally, as the method has a modular structure, it can be easily extended with data on new locations, context, or target populations, making it transferable to vaccination, chronic disease management, maternal health, and emergency response.^38^

This study has several limitations that warrant consideration. Its focus on the HCV-specific initiative may constrain generalizability to broader primary care or other healthcare contexts. The analysis, conducted within South Carolina’s Upstate region, may require validation in diverse geographic settings, such as coastal areas or mountainous areas, to assess its broader applicability. Demand was modeled using residential centroids weighted by uninsured population counts, overlooking daytime population movements, which may affect urban utilization patterns. Additionally, the analysis did not capture longitudinal or seasonal variations in MHC use. Moreover, this study did not consider the current hospital location or pharmacy location while placing the MHC. The model considered driving and walking separately and did not consider public transit networks in urban areas. Future research could enhance the model by integrating all modal networks, current location of hospitals or pharmacies or clinics, real-time mobility data, patient preferences to refine its responsiveness and impact. As we continue collaborating with CRH to inform future MHC deployments, there is a strong opportunity to validate this model prospectively, assessing whether real-world placements based on these recommendations align with projected service outcomes. Most importantly, while the proposed framework identifies optimal locations based on demand and access metrics, it does not guarantee that model-selected sites are operationally feasible for MHC deployment where factors such as permitting,^39^ timing,^40^ or logistical constraints^41^ may limit actual implementation.

However, the most significant limitation of this analytical approach is its focus solely on spatial accessibility, which, while necessary, is not sufficient to guarantee utilization. Our own findings suggest this: the attenuated effect of geographic access on uptake in urban settings indicates that analytical strategies for deployment may not yield their full potential without concurrent efforts to build community trust and partnerships.^7,32–35^ The model can identify the optimal placement, but cannot secure the trust and partnership required for real-world adoption. This is also vital in rural and historically marginalized communities, where distrust of outside institutions can be high.^43^ Therefore, the proposed framework should be viewed not as a replacement for community engagement, but as its complement. The data-driven site recommendations provide a powerful support tool for community partners, who can then co-design services through trusted local organizations while tracking metrics to learn and scale.^44–46^

In conclusion, this study offers a replicable, policy-ready framework for MHC site selection that strengthens spatial accessibility and operational efficiency. By replacing convenience-based practices with systematic, data-driven planning that integrates fine-scale geospatial analytics with real-world implementation, this framework expands access for uninsured and underserved groups while establishing a scalable basis for mobile healthcare delivery.

## Contributors

ST and LR conceptualized this study. ST conducted all data analyses and visualization in this study, with statistical analysis supervised by LR. ST, DW, BW, and LR developed the methodology for the study. ST wrote the original draft. LR performed project administration and funding. All authors interpreted study findings. All authors contributed to reviewing and editing the work, had access to all the data, approved the final version, agreed to be accountable for all aspects, and had final responsibility for the decision to submit for publication.

## Data sharing

Community-level data is publicly available and can be accessed through the United States Census Bureau American Community Survey. Due to patient confidentiality, data from Clemson Rural Health’s HCV screening program cannot be shared.

## Acknowledgments

This study was supported by the National Library of Medicine of the National Institutes of Health (R01LM014193), the National Institute on Drug Abuse of the National Institutes of Health (R33DA059892), and the Center for Forecasting and Outbreak Analytics of the Centers for Disease Control and Prevention (NU38FT000011). The HCV screening and treatment program was supported by Gilead Sciences, Inc., (IN-US-987-5892) with additional funding specifically for uninsured patients from South Carolina Center for Rural and Primary Healthcare (2015593). The funders had no role in the design, conduct, reporting of the study, or decision to submit for publication. During the preparation of this work the authors used ChatGPT-5 (OpenAI) to improve readability and clarity in parts of the manuscript. After using this tool, the authors reviewed and edited the content as needed and take full responsibility for the content of the publication.

