## Supplemental Files for "Optimizing Mobile Health Clinic Placement via Geospatial Modeling"

### Supplementary Appendix


| Table of content | Title | Page |
|---|---|---|
| Supplementary Material 1 | Single site scenario: minimizing walking time | 1 |
| Supplementary Figure 1 | Location allocation for single site scenario for walking time | 1 |
| Supplementary Material 2 | Multi-site scenario: minimizing walking time | 2 |
| Supplementary Figure 2 | Location allocation for multi-site scenario for walking time | 2 |
| Supplementary Table 1 | Relative and Absolute Gain in Urban and Rural Settings | 3 |
| Supplementary Figure 3 | Per-site service area uninsured population coverage increases | 3 |
| Supplementary Table 2 | Estimated effects on MHC visits (utilizations) based on 5-minute drive and 10-minute walk service areas. | 4 |
| Supplementary Table 3 | For Case Study 1, Population Coverage and Site Rankings For 20 Proposed MHC Sites by Uninsured Adults Aged 35–65 Within 5-Minute Drive and 10-Minute Walk Service Areas. | 5 |
| Supplementary Table 4 | For Case Study 2, Population Coverage and Site Rankings For 32 MHC Sites by Uninsured Adults Aged 35–65 within 10-Minute Drive and 10-Minute Walk Service Areas, with ZIP Code Mortality Rates. | 6 |
| Supplementary Table 5 | For Case Study 3, Population Coverage and Site Rankings for Six Candidate MHC Sites by Uninsured Adults Aged 35–65 within 5-Minute Drive and 10-Minute Walk Service Areas. | 8 |

**Supplementary Material 1: Single site scenario: minimizing walking time**

For ZIP code 29611, an urban area near Greenville, Figure (Supplementary figure 1) illustrates the spatial distribution of the observed MHC site, the proposed site selected from 107 candidate sites for walking mode. The proposed site serves 76 census blocks within a 10-minute walking service area, yielding an average walking time of 7.06 minutes per person. The proposed site offers improvement over the observed location in terms of proximity to more uninsured population as it is situated closer to the densest cluster of census blocks. The service area analysis shows the observed MHC site served just 6 uninsured individuals within a 10-minute walk, while the proposed site serves 974, an approximate 16-fold increase.

**Supplementary Figure 1: Location allocation for single site scenario for walking time**

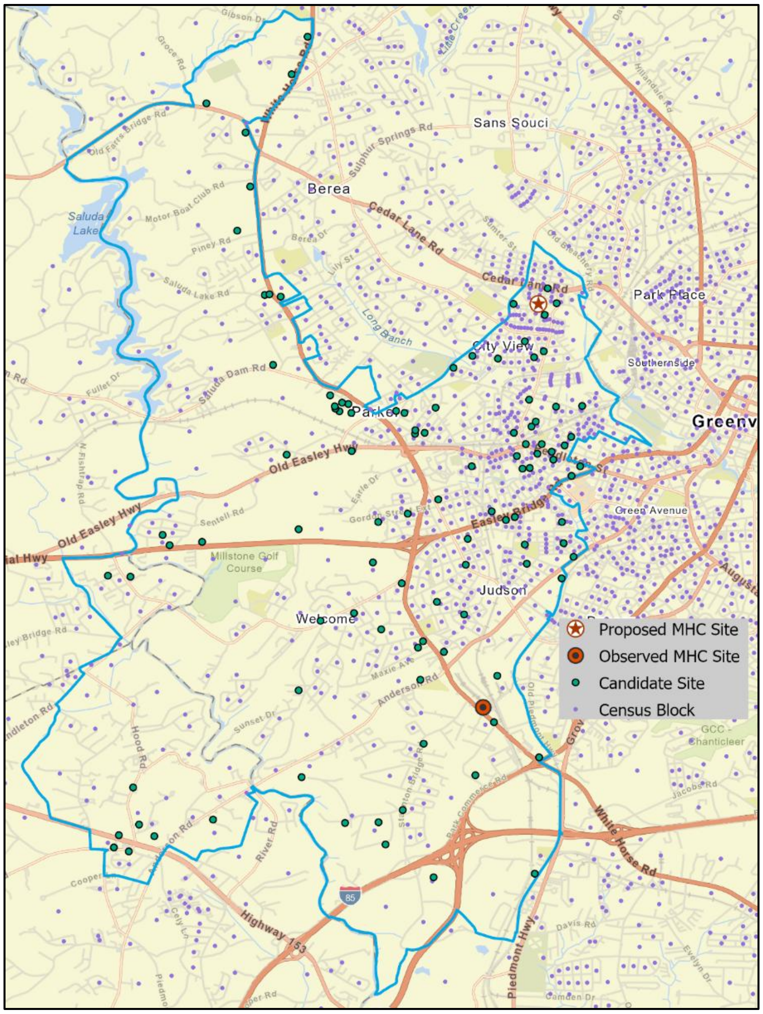


Note: Location allocation for single site scenario for walking time showing spatial distribution of proposed site, observed site, candidate sites, and census block centroids in an urban ZIP code.

**Supplementary Material 2: Multi-site scenario: minimizing walking time**

For ZIP code 29678, a rural area near Seneca, five sites proposed from 72 potential locations (Figure B2). The proposed sites collectively serve 140 census blocks within a 10-minute walking distance, resulting in an average walking time of 7.98 minutes per person. The map shows that the proposed sites are more spatially dispersed across the ZIP code, avoiding the clustering seen in observed sites. This reduces the overlap of the service area and improves coverage efficiency. Based on service area analysis, the proposed MHC sites serve 582 unique uninsured individuals within 10-minute walking areas, up from 248 previously, representing a 135% increase.

**Supplementary Figure 2: Location allocation for multi-site scenario for walking time**

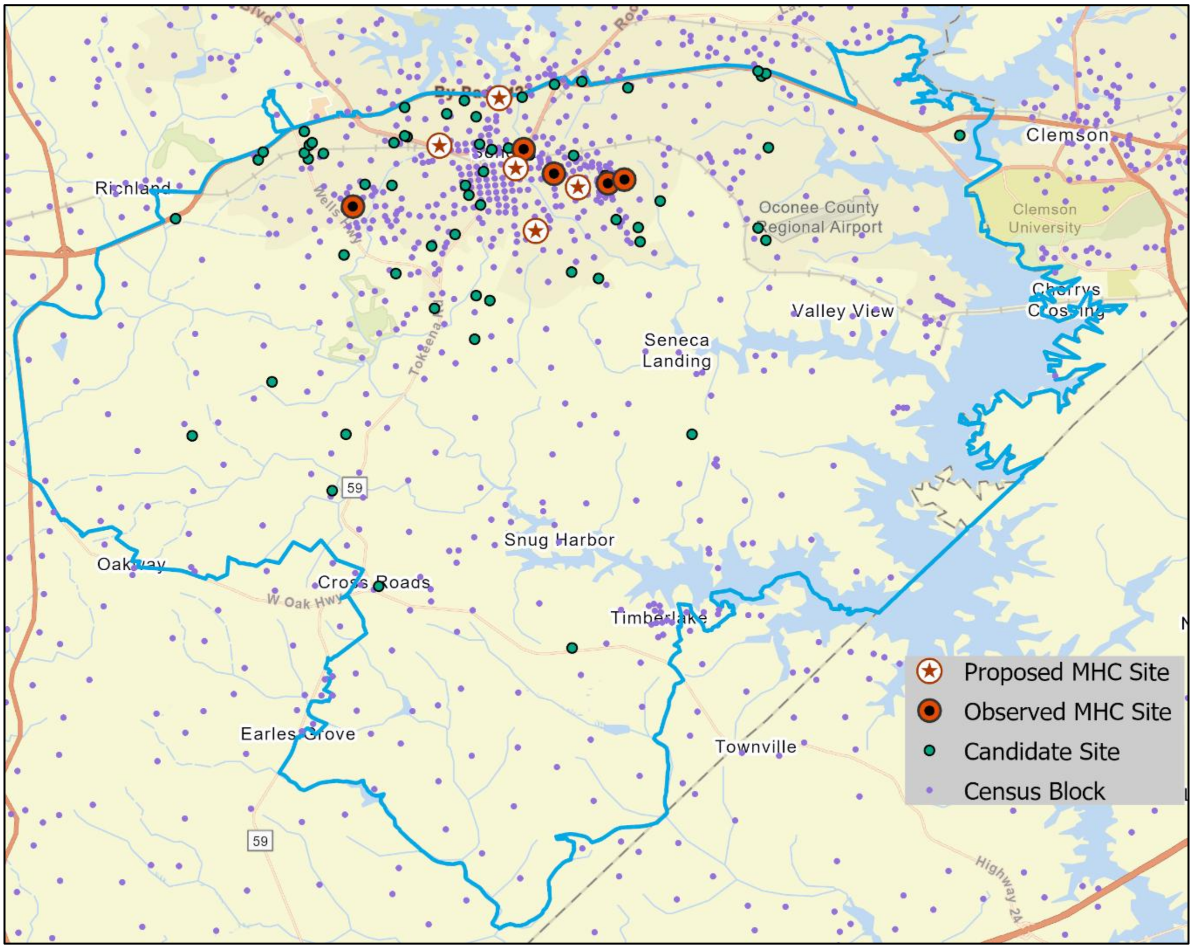


Note: Location allocation for multi-site scenario for walking time showing spatial distribution of three proposed sites, three observed sites, candidate sites, and census block centroids in a rural ZIP code.

**Supplementary Table 1: Relative and Absolute Gain in Urban and Rural Settings**

| **Setting** | **Uninsured population in driving service area** | | **Abs Δ** | **% Δ** | **Uninsured population in walking service area** | | **Abs Δ** | **% Δ** |
|---|---|---|---|---|---|---|---|---|
| | Observed Sites | Proposed Sites | | | Observed Sites | Proposed Sites | | |
| **Urban (8 ZIPs; 11 sites)** | 13,023 | 28,148 | 15,125 | 116% | 1,740 | 3,867 | 2,127 | 122% |
| **Rural (12 ZIPs; 17 sites)** | 9,020 | 13,810 | 4,790 | 53% | 900 | 2,333 | 1,433 | 159% |

**Supplementary Table 2: Estimated effects on MHC visits (utilizations) based on 5-minute drive and 10-minute walk service areas.**

| | **5 min drive service area** | | | **10 min walk service area** | | |
|---|---|---|---|---|---|---|
| | Estimate | Std. Error | P-value | Estimate | Std. Error | P-value |
| **Uninsured Population** | 0.0093 | 0.0032 | 0.0037 | 0.2575 | 0.0993 | 0.0095 |
| **Urban Area** | 1.5997 | 0.6865 | 0.0198 | 1.2186 | 0.4951 | 0.0139 |
| **Uninsured × Urban (Interaction)** | -0.0092 | 0.0032 | 0.0038 | -0.2605 | 0.0994 | 0.0088 |

**Supplementary Table 3: For Case Study 1, Population Coverage and Site Rankings For 20 Proposed MHC Sites by Uninsured Adults Aged 35–65 Within 5-Minute Drive and 10-Minute Walk Service Areas.**

| | Service area 5 m drive | | | Service area 10 m walk | | | Combined | |
|---|---|---|---|---|---|---|---|---|
| **Site ID** | **Pop** | **No_HI_35-65** | **Rank** | **Pop** | **No_HI_35-65** | **Rank** | **No_HI_35-65** | **Rank** |
| **S-1** | 5893 | 566 | 1 | 455 | 52 | 5 | 618 | 1 |
| **S-2** | 5709 | 464 | 2 | 314 | 10 | 9 | 474 | 4 |
| **S-3** | 3262 | 459 | 3 | 403 | 145 | 1 | 604 | 2 |
| **S-4** | 5959 | 455 | 4 | 992 | 141 | 2 | 596 | 3 |
| **S-5** | 1434 | 325 | 5 | 327 | 127 | 3 | 452 | 5 |
| **S-6** | 4826 | 307 | 6 | 366 | 18 | 7 | 325 | 7 |
| **S-7** | 1064 | 304 | 7 | 30 | 12 | 8 | 316 | 8 |
| **S-8** | 3400 | 291 | 8 | 46 | 6 | 10 | 297 | 9 |
| **S-9** | 1032 | 287 | 9 | 161 | 62 | 4 | 349 | 6 |
| **S-10** | 2994 | 228 | 10 | 184 | 23 | 6 | 251 | 10 |
| **S-11** | 683 | 194 | 11 | 0 | 0 | 16 | 194 | 11 |
| **S-12** | 1983 | 121 | 12 | 0 | 0 | 16 | 121 | 12 |
| **S-13** | 1652 | 98 | 13 | 4 | 0 | 15 | 98 | 13 |
| **S-14** | 2748 | 72 | 14 | 152 | 5 | 11 | 77 | 14 |
| **S-15** | 2956 | 69 | 15 | 65 | 1 | 13 | 70 | 15 |
| **S-16** | 216 | 33 | 16 | 0 | 0 | 16 | 33 | 16 |
| **S-17** | 166 | 22 | 17 | 0 | 0 | 16 | 22 | 17 |
| **S-18** | 144 | 22 | 18 | 0 | 0 | 16 | 22 | 17 |
| **S-19** | 161 | 11 | 19 | 25 | 2 | 12 | 13 | 19 |
| **S-20** | 370 | 7 | 20 | 26 | 1 | 14 | 8 | 20 |

*Notes:*
*Pop: Number of populations in the service area.*
*No_HI_35-65: Number of people aged 35 to 65 without health insurance in the service area.*
*Rank: Rank based on No_HI_35-65.*
*Table sorted based on combined ranking.*
*Site names are deidentified.*

**Supplementary Table 4: For Case Study 2, Population Coverage and Site Rankings For 32 MHC Sites by Uninsured Adults Aged 35–65 within 10-Minute Drive and 10-Minute Walk Service Areas, with ZIP Code Mortality Rates.**

| Site ID | 10-Min Drive No_HI_35-65 | 10-Min Drive Rank | 10-Min Walk No_HI_35-65 | 10-Min Walk Rank | Combined No_HI_35-65 | Combined Rank | Zip Code | County | Deaths | Death Rate (per 1,000) | Rank (Death rate) |
|---|---|---|---|---|---|---|---|---|---|---|---|
| **S-1** | 4,846 | 1 | 184 | 1 | 5,030 | 1 | Z-3 | C-1 | 139 | 3.5 | 17 |
| **S-2** | 4,072 | 2 | 4 | 25 | 4,076 | 2 | Z-3 | C-1 | 139 | 3.5 | 17 |
| **S-3** | 3,263 | 4 | 173 | 2 | 3,436 | 3 | Z-5 | C-3 | 110 | 7.7 | 2 |
| **S-4** | 3,267 | 3 | 25 | 19 | 3,292 | 4 | Z-9 | C-1 | 148 | 3.5 | 17 |
| **S-5** | 3,147 | 5 | 104 | 6 | 3,251 | 5 | Z-5 | C-3 | 110 | 7.7 | 2 |
| **S-6** | 3,004 | 7 | 90 | 7 | 3,094 | 6 | Z-5 | C-3 | 110 | 7.7 | 2 |
| **S-7** | 3,021 | 6 | 0 | 30 | 3,021 | 7 | Z-6 | C-5 | 154 | 4.8 | 13 |
| **S-8** | 2,823 | 8 | 73 | 9 | 2,896 | 8 | Z-8 | C-1 | 196 | 3.8 | 16 |
| **S-9** | 2,017 | 10 | 124 | 5 | 2,141 | 9 | Z-7 | C-4 | 130 | 4.7 | 14 |
| **S-10** | 1,942 | 12 | 147 | 3 | 2,089 | 10 | Z-7 | C-4 | 130 | 4.7 | 14 |
| **S-11** | 1,915 | 16 | 145 | 4 | 2,060 | 11 | Z-7 | C-4 | 130 | 4.7 | 14 |
| **S-12** | 2,040 | 9 | 0 | 30 | 2,040 | 12 | Z-10 | C-1 | 157 | 2.4 | 19 |
| **S-13** | 1,920 | 15 | 82 | 8 | 2,002 | 13 | Z-7 | C-4 | 130 | 4.7 | 14 |
| **S-14** | 1,985 | 11 | 9 | 22 | 1,994 | 14 | Z-10 | C-1 | 157 | 2.4 | 19 |
| **S-15** | 1,936 | 13 | 43 | 12 | 1,979 | 15 | Z-7 | C-4 | 130 | 4.7 | 14 |
| **S-16** | 1,926 | 14 | 29 | 15 | 1,955 | 16 | Z-3 | C-1 | 139 | 3.5 | 17 |
| **S-17** | 1,843 | 17 | 47 | 11 | 1,890 | 17 | Z-7 | C-4 | 130 | 4.7 | 14 |
| **S-18** | 1,835 | 18 | 29 | 16 | 1,864 | 18 | Z-10 | C-1 | 157 | 2.4 | 19 |
| **S-19** | 1,661 | 19 | 29 | 17 | 1,690 | 19 | Z-7 | C-4 | 130 | 4.7 | 14 |
| **S-20** | 1,541 | 22 | 57 | 10 | 1,598 | 20 | Z-6 | C-5 | 154 | 4.8 | 13 |
| **S-21** | 1,542 | 20 | 43 | 13 | 1,585 | 21 | Z-6 | C-5 | 154 | 4.8 | 13 |
| **S-22** | 1,542 | 21 | 31 | 14 | 1,573 | 22 | Z-6 | C-5 | 154 | 4.8 | 13 |
| **S-23** | 1,422 | 24 | 25 | 20 | 1,447 | 23 | Z-1 | C-2 | 135 | 6.6 | 5 |
| **S-24** | 1,436 | 23 | 8 | 23 | 1,444 | 24 | Z-2 | C-2 | 120 | 6.2 | 10 |
| **S-25** | 1,404 | 25 | 28 | 18 | 1,432 | 25 | Z-1 | C-2 | 135 | 6.6 | 5 |
| **S-26** | 1,368 | 26 | 5 | 24 | 1,373 | 26 | Z-4 | C-3 | 271 | 6.1 | 11 |
| **S-27** | 1,342 | 27 | 13 | 21 | 1,355 | 27 | Z-4 | C-3 | 271 | 6.1 | 11 |

| | | | | | | | | | | | |
|---|---|---|---|---|---|---|---|---|---|---|---|
| **S-28** | 1,300 | 28 | 3 | 27 | 1,303 | 28 | Z-1 | C-2 | 135 | 6.6 | 5 |
| **S-29** | 812 | 29 | 3 | 26 | 815 | 29 | Z-1 | C-2 | 135 | 6.6 | 5 |
| **S-30** | 618 | 30 | 1 | 29 | 619 | 30 | Z-6 | C-5 | 154 | 4.8 | 13 |
| **S-31** | 572 | 31 | 0 | 30 | 572 | 31 | Z-2 | C-2 | 120 | 6.2 | 10 |
| **S-32** | 424 | 32 | 1 | 28 | 425 | 32 | Z-11 | C-3 | 50 | 6.5 | 8 |

*Notes:*
*No_HI_35-65: Number of uninsured individuals aged 35–65.*
*Rank: Based on No_HI_35-65 in each category.*
*The death rate is for respiratory diseases, including COVID-19, influenza, and RSV, for the years 2020-2022.*
*Rank for death and death rate are based on the ZIP code. Some of the ranked ZIPs are not here, as there were no current sites in those ZIP. For example, the top-ranked ZIP code for death rate.*
*Table sorted based on combined ranking.*
*Site names, ZIP Code, and County Names are deidentified.*

**Supplementary Table 5: For Case Study 3, Population Coverage and Site Rankings for Six Candidate MHC Sites by Uninsured Adults Aged 35–65 within 5-Minute Drive and 10-Minute Walk Service Areas.**

| Site ID | 5-Min Drive No_HI_35-65 | 5-Min Drive Rank | 10-Min Walk No_HI_35-65 | 10-Min Walk Rank | Combined No_HI_35-65 | Combined Rank |
|---|---|---|---|---|---|---|
| **S-1** | 506 | 2 | 88 | 1 | 594 | 1 |
| **S-2*** | 533 | 1 | 18 | 2 | 551 | 2 |
| **S-3** | 404 | 3 | 1 | 5 | 405 | 3 |
| **S-4*** | 327 | 4 | 0 | 6 | 327 | 4 |
| **S-5*** | 287 | 5 | 6 | 4 | 293 | 5 |
| **S-6** | 204 | 6 | 11 | 3 | 215 | 6 |

*Notes:*
*No_HI_35-65: Number of uninsured individuals aged 35–65.*
*Rank: Based on No_HI_35-65 in each category.*
*Site names are deidentified.*
*Table sorted based on combined ranking.*
*The top three choices for serving uninsured people aged 35-64 are: S-1, S-2, and S-3.*
**S-2, S-4, and S-5 sites have overlapping 5-minute drive service areas. Therefore, if one of these sites is selected, the others should not be included when choosing 3 out of the 6 available sites.*
